# Cognitive improvement after dual-task training in Parkinson’s disease: a follow-up study

**DOI:** 10.1101/2022.04.27.22274290

**Authors:** Dalma Szögedi, Trevor W. Stone, Elek Dinya, Judit Málly

**Author notes:** Corresponding Author (MD, PhD, Habil.). T W Stone, Kennedy Institute of Rheumatology, University of Oxford.

## Abstract

The reaction times of patients with Parkinson’s disease (PD) in tests of simultaneous dual-task accuracy are dependent on the subjects’ cognitive ability. We now report that one week training on dual-task tests improved cognitive function. Forty-six PD patients were compared with 47 age matched healthy controls and 26 patients were followed for one year. Five dual-task tests consisting of a primary cognitive task to be performed simultaneously with a secondary motor task were repeated for five consecutive days. Testing was repeated after 6 and 12 months. Participants’ reaction times, including Hits and Misses, were quantified. Initial tests indicated slower reaction times in patients compared to controls, with fewer Hits and more Misses, especially in PD patients over 65 years of age. Training by daily repeated dual-task tests improved performance within 3 days (p < 0. 01 or p < 0.001), with no deterioration after 6 months. We conclude that dual-tasks are objective and sensitive tests for detecting early cognitive difficulties in PD, with improvements in both by repeated exposure to testing. A few days of testing produced cognitive improvement lasting many months. It is recommended that the use of simultaneous, dual-task testing is used to produce long-lasting improvement of cognitive function in PD patients.

## Introduction

Few interventions, pharmacological or non-pharmacological, have a marked effect on cognitive decline in Parkinson’s disease (PD) [1-3] although recent studies with dual task training have reported improvements in attention and executive function in PD [4 -7]. The performance of simultaneous dual-task tests assesses changes in the reaction time of patients when a primary task is associated with a second, unrelated [8 -11]. In particular, cognitive tasks influence various parameters of walking [12 - 15] or other motor activity [16] in PD patients. Dual-task performance assessed by interactive computer applications can evaluate a wide range of executive cognitive activities in PD with Hoehn-Yahr (HY) stages II and III symptoms [17]. Cognitive status showed a high correlation with dual-task performance in patients with mild cognitive impairment (MCI) detected by auditory Stroop test. Reaction times were slower in patients with cognitive impairment, although they did not correlate with the motor status of PD patients assessed by UPDRS –III. [18]. This statement is based on PD patients with Hoehn-Yahr (HY) stages II and III, between which there is a substantial loss of motor function and balance, but we considered that it might be possible to show changes between stages H-Y I-II in which attention and executive functions are also compromised. Therefore, the objective of this study was to determine whether cognitive decline in the early stages of PD can be assessed by dual-task performance. Although dual task testing provides only an overall, global assessment of cognition function, it was felt adequate for an initial study here. Previous favorable results have used dual-task training twice a week for several weeks [4 - 7].

We have now examined shorter periods of training for only a few days, but using several forms of dual-tasking. We also determined the degree of cognitive deterioration after 6 and 12 months.

## Materials and methods

### Ethics

The Regional Ethics Committee of the Petz Aladár County Hospital in Győr, Hungary, provided permission for the present trial (The number of permission: 76-1-6/2019). The subjects gave their written informed consent at the onset of the trials, according to the Helsinki Declaration. The study was listed on the ISRCTN registry with study ID ISRCTN49538525 (www-isrctn.com/ISRCTN49538525).

### Participants

Patients with PD (N = 46) were compared with 47 age-matched healthy controls. The inclusion criteria were as follows:- (a) the presence of Parkinson’s disease responding well to levodopa; (b) no evidence of dementia or minimal mental impairment according to different cognitive tests (c) no other chronic disease. Patients with PD in H-Y I and II were included in this study. The subjects were divided into two groups according to their age (under and over 65 years) (Table 1). Patients were given levodopa retard and entacapone. Patients showed a mixture of slight tremor and hypokinesis without any functional impairment.

**Table 1.** The demographic data of the comparative study with dual-task tests

|  | Control | Parkinson's | Control | Parkinson's |
| --- | --- | --- | --- | --- |
| | $\leq 65$ years | | $> 65$ years | |
| Number | 29 | 26 | 18 | 20 |
| Age (yrs) | $54.6 \pm 8.8$ | $58.3 \pm 7.1$ | $68.1 \pm 2.6$ | $71.5 \pm 4.1$ |
| Female; male | 16; 13 | 14; 12 | 10; 8 | 9; 11 |
| Duration of disease (yrs) | | $5.6 \pm 3.0$ | | $5.8 \pm 3.5$ |
| H-Y stages | | $1.5 \pm 0.55$ | | $1.7 \pm 0.58$ |
| UPDRS total. | | $21.94 \pm 11.46$ | | $23.2 \pm 7.1$ |
| T/H |  | 15/11 |  | 11/9 |
| Dose of levodopa (mg/d) | | $357.8 \pm 161.8$ | | $320.5 \pm 182.0$ |

The table 1 shows the demographic data. The controls and the patients with Parkinson’s disease were divided into two groups according their ages (≤ 65 years and > 65 years). There was no significant difference between their duration of disease (p = 0.9110), dose of levodopa (p = 0.3805), total score of UPDRS (p = 0.6339), Hoehn-Yahr stage (H-Y stage) (p = 0.2766). T = tremor H = hypokinezis

Twenty-six patients with PD (N = 13 ≤ 65 years, N = 13 > 65 years) were followed for one year. The reduction in the number of patients compared to the initial number was due to the distance of their habitation. Their severity in UPDRS total between the initial group (I) ≤ 65 years and followed patients (F) ≤ 65 years (p = 0.4681) and H-Y stages in I group ≤ 65 years and F group ≤ 65 years (p = 0.3308) did not differ significantly from each other. Above 65 years the significance in UPDRS total (p = 0.7366) and in H-Y (p= 0.6545) was. Their detailed demographic data are involved in Table 2.

**Table 2.** Demographic data of patients with Parkinson’s Disease included in this follow-up study.

|  | **$\leq 65$ YEARS PD** |  |  | **$> 65$ YEARS PD** |  |  |
| --- | --- | --- | --- | --- | --- | --- |
|  | Baseline | Half year | One year | Baseline | Half year | One year |
| Number of patients | 13 | 13 | 13 | 13 | 13 | 13 |
| Age | $57.6 \pm 7.5$ | | | $71.3 \pm 3.5$ | | |
| Duration of the disease | $4.8 \pm 2.7$ | | | $5.6 \pm 3.6$ | | |
| UPDRS total | $18.9 \pm 11.0$ | $20.3 \pm 12.9$ | $21.0 \pm 11.4$ | $21.3 \pm 7.3$ | $20.4 \pm 10.5$ | $21.4 \pm 9.6$ |
| H-Y stages | $1.38 \pm 0.5$ | $2.00 \pm 0.7$ | $1.96 \pm 0.66$ | $1.65 \pm 0.55$ | $1.69 \pm 0.59$ | $1.80 \pm 0.63$ |
| T/H | 7/6 |  |  | 6/7 |  |  |
| Dose of levotopa (mg/day) | $315.3 \pm 158.6$ | $376.9 \pm 183.2$ | $369.2 \pm 201.5$ | $269.2 \pm 163.9$ | $284.6 \pm 189.7$ | $288.4 \pm 188.3$ |

The table 2 shows the demographic data of patients with Parkinson’s disease (PD) (N = 26). Comparing patients ≤ 65 years and > 65 years in the categories of “Baseline”, “Half year” and “One year” time points, there were no significant differences in the duration of disease (p = 0.5302), total score of Unified Parkinson Disability Rating Scale (UPDRS) (p = 0.5719; 0.9708; 0.9272), the dose of levodopa (p = 0.5203; 0.2018; 0.2629) and Hoehn-Yahr stages (H-Y stages) (p = 0.2676; 0.2059; 0.5248). T = tremor H = hypokinezis

### Study Design

The first part of the present work was a comparative study with age-matched healthy controls. The second part of the follow-up research was a self-controlled experiment with PD patients.

### Assessments

Dual-tasks performances were examined using Dividat Senso equipment (HUR, Finland), with subjects standing on a glass platform overlying 20 force sensors. A cognitive task was combined with a motor activity task in each test period, with subjects asked to focus on a game presented on a visual monitor. For the motor task, patients were asked to detect an object appearing at one edge (top, bottom, right or left) of the screen and were required to react immediately using leg movements. Continuously focusing on the cognitive task (attention and decision), the leg movements on both sides were controlled by the person. Five dual-task tests were applied.

For cognitive testing, at first a ‘Bird’ task was used in which a bird had to be selected from different colored figures. In a ‘Simple’ task, red spots were shown at different positions. In the game ‘Divided’, red spots were linked with high and low-pitched sounds, and the participants were required to make goal directed movements. In the game ‘Habitat’ four different animals had to be allocated to their appropriate living area. When they were at a wrong place, persons had to make a step. Dual-task interactions were quantified by the average reaction times. In the game of ‘Target’, black bullets moved around the monitor, with different speeds, and the subject was asked to make a step, when the black bullet touched the target. Correct and incorrect responses were recorded. The tasks lasted for one and a half minutes and were repeated each day for five consecutive days. The computer gave the average performance in less than a minute and half. The training with these dual-task performances was replicated at 6 and 12 months.

The following traditional tests were also applied; the Mini Mental Rating Scale [19], the Ziehen Ranschburg Word Pair Test, Trail Making Test [20], Clock Drawing Test [21], and the Hamilton Depression Scale Tests [22]. For the detection and quantitation of Parkinsonian symptoms the Hoehn-Yahr Stages were used [23] together with the Unified Parkinson Disability Rating Scale [24]. Walking ability was measured as distance walked in 6 mins (in m), and time taken to walk 10m (in sec). The walking tests were performed on the first and fifth days of training. The cognitive test was administered by a psychologist, the dual-task test with Dividat Senso were controlled by a physiotherapist and the mathematical statistical analysis was done by a mathematician.

### Statistical analysis

Results are expressed as the mean ± standard deviation of mean and sample size for each age group with Parkinson. The normality of data was checked by applying the Shapiro-Wilk’s test and the homogeneity of variances was assessed through the Levene’s test. For baseline values, we performed the necessary statistical analysis with the nonparametric Mann-Whitney test to determine significant differences for the PD age groups examined (<65,> 65 years), but no significant differences were found. The means of different date (baseline, half year, one year data) were compared by nonparametric Friedman ANOVA, significance values have been adjusted by the Bonferroni correction for multiple tests. The analysis was two sided with a level of significance of α = 0.05. All statistical analyses were performed using the SAS 9.4 (SAS Institute Inc., Cary, NC, USA) software package.

## Results

There was no difference in symptom severity or cognitive performance in PD patients under or over 65 years of age assessed by UPDRS total and H-Y stages (Table 1). However, there was a significant difference between the age matched healthy controls and the patients with PD ≤ 65 years in their dual-task tests of Bird (p < 0.001), Divided (p < 0.05) and Misses (p < 0.05) (Table 3).

**Table 3.** Comparing the patients’ and controls’ results in the dual-task activity during training with dual-task tests ≤ 65 years.

|  | ≤ 65 Years |  |  |  |  |  |
| --- | --- | --- | --- | --- | --- | --- |
|  | Controls |  |  | Parkinson's disease |  |  |
|  | N = 29 |  |  | N = 26 |  |  |
|  | DAYS |  |  |  |  |  |
|  | 1<sup>st</sup> | 3<sup>rd</sup> | 5<sup>th</sup> | 1<sup>st</sup> | 3<sup>rd</sup> | 5<sup>th</sup> |
| Bird | 1.30 ± 0.20 | 1.10 ± 0.12 <sup>+++</sup> | 1.02 ± 0.11 <sup>+++</sup> | 1.75 ± 0.69 <sup>***</sup> | 1.37 ± 0.37 <sup>+++ **</sup> | 1.31 ± 0.32 <sup>+++ **</sup> |
| Simple | 0.94 ± 0.15 | 0.83 ± 0.30 | 0.78 ± 0.09 | 1.31 ± 0.69 | 1.04 ± 0.37 <sup>+++</sup> | 0.98 ± 0.23 <sup>+++</sup> |
| Divided | 1.01 ± 0.17 | 0.94 ± 0.12 <sup>++</sup> | 0.88 ± 0.11 <sup>+++</sup> | 1.18 ± 0.45<sup>*</sup> | 1.09 ± 0.31 | 0.07 ± 0.22 <sup>+ *</sup> |
| Habitat | 1.67 ± 0.21 | 1.50 ± 0.13 <sup>+++</sup> | 1.41 ± 0.12 <sup>+++</sup> | 1.78 ± 0.25 | 1.62 ± 0.30 <sup>++</sup> | 1.68 ± 0.27 <sup>++ ***</sup> |
| Hits | 84.10 ± 9.03 | 109.03 ± 17.13 <sup>+++</sup> | 119.93 ± 20.13 <sup>+++</sup> | 72.00 ± 24.00 | 86.80 ± 24.16 <sup>+++ **</sup> | 93.57 ± 27.46 <sup>+++ ***</sup> |
| Misses | 10.93 ± 7.25 | 10.17 ± 5.16 | 10.75 ± 6.82 | 17.30 ± 12.07<sup>*</sup> | 17.65 ± 9.28 <sup>**</sup> | 16.07 ± 10.50 <sup>*</sup> |

Table 3 shows the results of the dual-task performances of Parkinsonian patients ≤ 65 years compared with that of the controls. Data of Bird, Simple, Divided, Habitat are given in seconds of their mean ± S.D. Hits and Misses are given in the mean ± S.D. The symbol * means the significant difference between the controls and parkinsonian patients (* = p < 0.05, ** = p < 0.01, *** = p < 0.001). The symbol + means the significant difference between the results of first days and the values of the next days during training (+ = p < 0.05, ++ = p < 0.01, +++ = p < 0.001).

A significant difference was observed in the PD patients > 65 years in their initial five dual-task tests: Bird (p < 0.001), Simple (p < 0.001), Divided (p < 0.05), Hits (p < 0.05), Misses (p < 0.001) compared with the controls (Table 4).

**Table 4.**
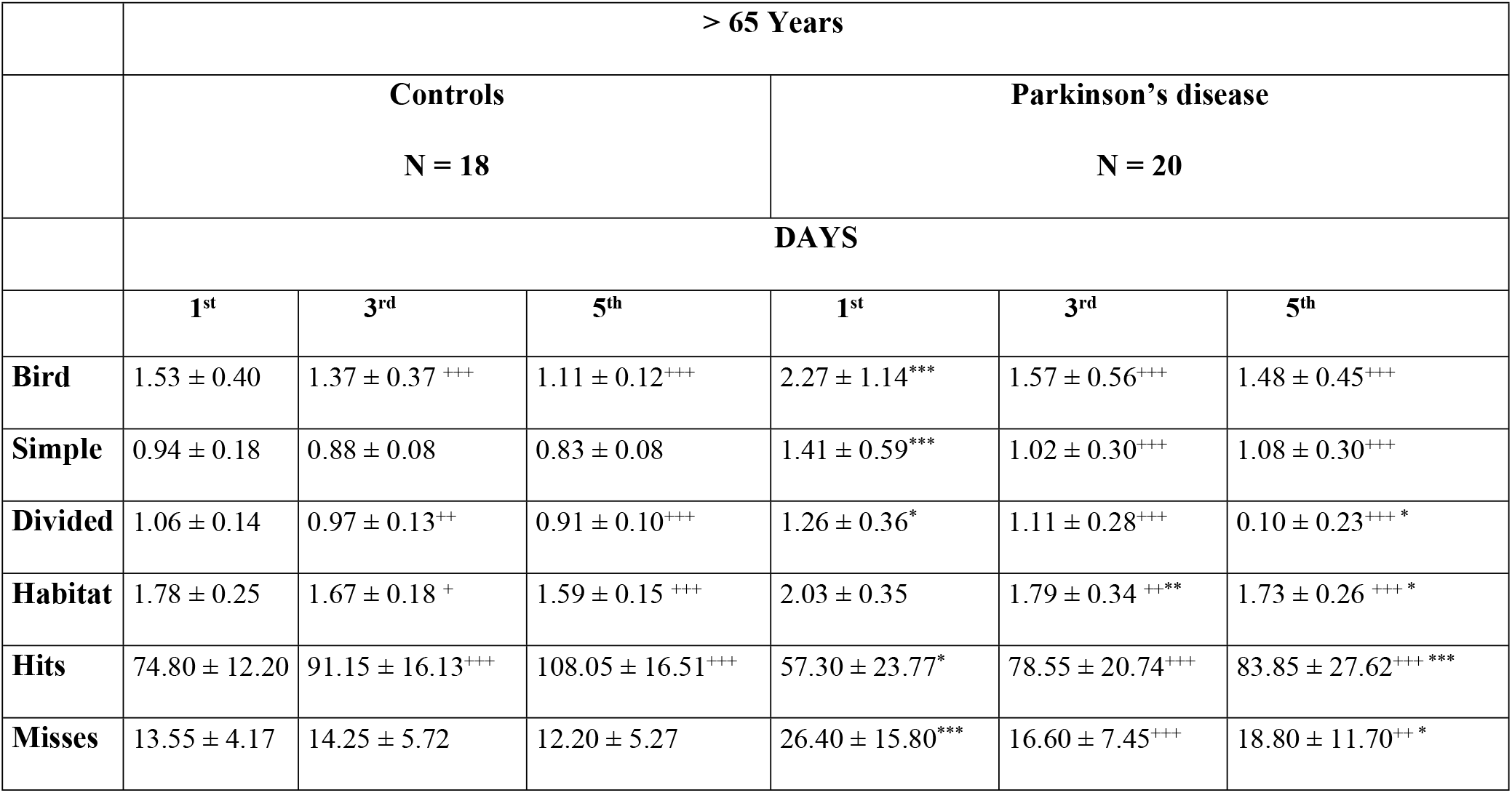
Comparing the patients’ and controls’ results in the dual-task activity during training with dual-task tests > 65 years.

Table 4 shows the results of the dual-task performances of Parkinsonian patients > 65 years compared with that of the controls. Data of Bird, Simple, Divided, Habitat are given in seconds of their mean ± S.D. Hits and Misses are given in the mean ± S.D.

The symbol * means the significant difference between the controls and parkinsonian patients (* = p < 0.05, ** = p < 0.01, *** = p < 0.001). The symbol + means the significant difference between the results of first days and the values of the next days during training (+ = p < 0.05, ++ = p < 0.01, +++ = p < 0.001).

The two Parkinsonian groups significantly differed from each other in the dual-task activities to the detriment of the older group of patients, namely: Bird (p < 0.05), Habitat (p < 0.05), Hits (p < 0.05), Misses (p < 0.05) (Table 3 and 4), although the controls exhibited age-related differences in the dual-task test of Bird (p < 0.05). Dual-task training effectively influenced the results of dual-task activities in controls and PD patients (Table 3, 4). Our data were analyzed in two ways, comparing the results on the first day to the 3^rd^ and the 5^th^ days (Table 3, 4). In controls ≤ 65 years, the reaction times on dual-task Bird (p < 0.001), Divided (p < 0.01) and Habitat (p < 0.001) were decreased, and the Hits elevated (p <0.001), but the Simple and Misses tests did not change on the 3^rd^ day of training compared to the results on the 1^st^ day (Table 3). A similar improvement was detected on the 5^th^ day of training. PD patients ≤ 65 years showed reduced reaction times in Bird (p < 0.001), Simple (p ≤ 0.001) and Habitat (p < 0.01), but an elevation in Hits (p < 0.001) when results were taken on the 1^st^ day, but the Divided and Misses tests showed no alteration during the training. Similar favorable changes were also obtained on the 5^th^ day. Significant changes were noted with results on the 1^st^ day, although they did not reach control values. The patients’ results on day 5 were significantly different from the controls, in the Bird (p ≤ 0.01), Divided (p < 0.05), Habitat (p < 0.001), Hits (p < 0.001) and Misses (p < 0.05) paradigms (Table 3).

Comparable results were obtained in participants > 65 years, but the amount of change during dual-task training was more impressive than in the age matched group under 65 years. The controls > 65 years showed decreased reaction times in dual-task performances; Bird (p < 0.001), Divided (p < 0.01), Habitat (p <0.05), with an increase in the number of Hits (p < 0.001) on the 3^rd^ day of training. The changes were preserved on the 5^th^ day. The dual-task tests Simple and Misses did not change as in the younger control group of participants. The greatest changes were observed in the Parkinsonian patients > 65 years. All dual-task results were changed on the 3^rd^ day compared to the 1^st^ day (Table 4), and these changes further increased on the 5^th^ day after training (p < 0.001). However, the values of PD patients > 65 years remained elevated compared to the controls, namely: Divided (p < 0.05), Habitat (p < 0.05), number of Hits (p < 0.001) on the 5^th^ day of training. Even the number of Misses decreased in this group of patients, but at the end of training the results were significantly higher than that of the controls (p < 0.05).

### Repetition at 6 and 12 months

The short-term dual-task training was repeated after 6 and 12 months. After 6 months, average reaction times on the first day were lower than the baseline, but only the reaction times of ‘Bird’ (p < 0.05) and ‘Simple’ (p < 0.01) were decreased significantly. After one year these changes further increased on the first day of training (p < 0.001) compared to the baseline (Fig 1). The number of ‘Hits’ was elevated (Baseline (B): 85.1 ±16.1; Half year (H): 99.5 ±16.5 p < 0.05; one year (O): 110.7 ± 11.3 p < 0.001) in PD patients ≤ 65 years. No changes were noted in the 5-day dual-task tests of ‘Habitat’ and ‘Misses’.

**Fig 1.**
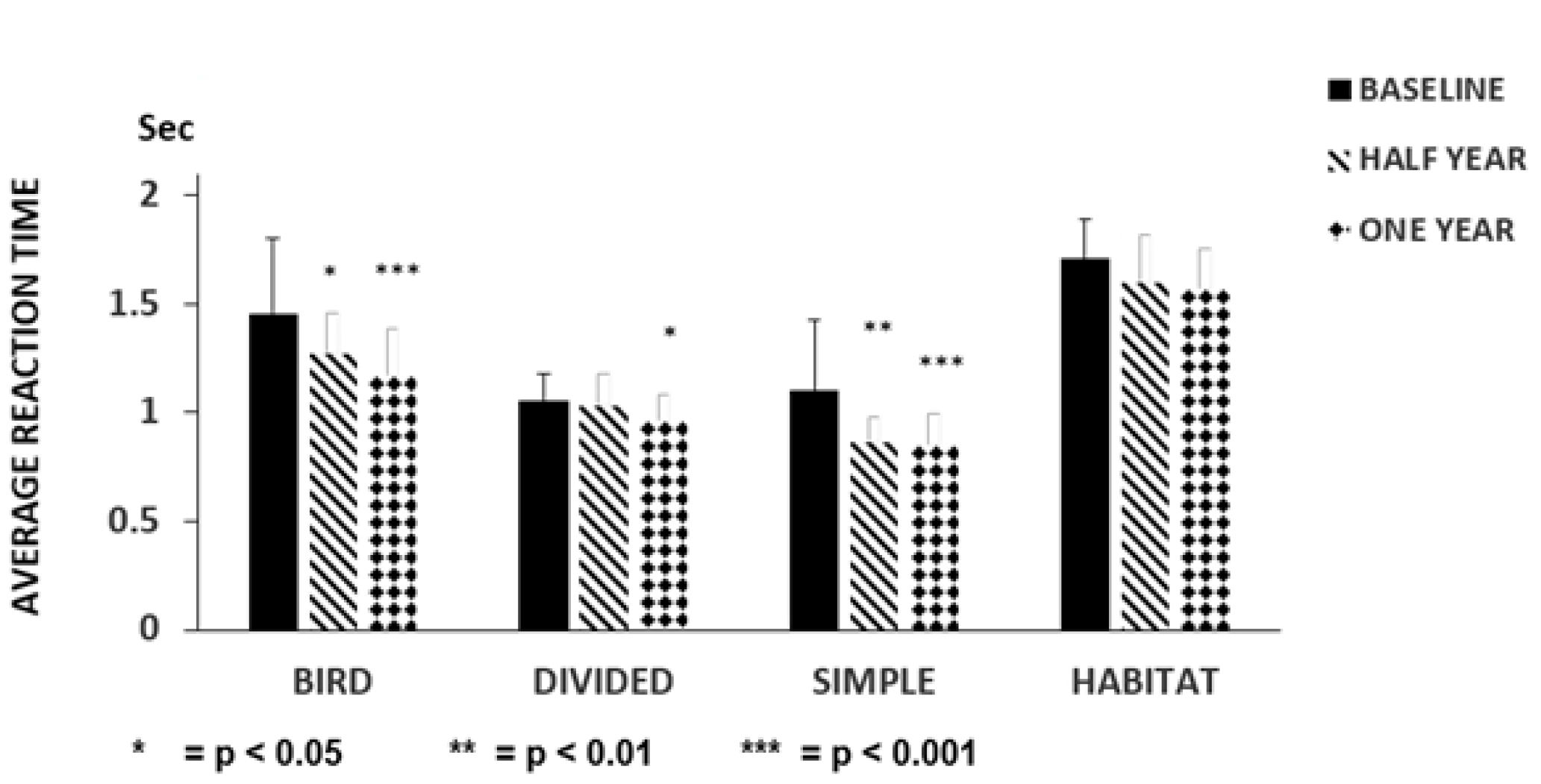
Follow-up of the effect of training with dual-task performances in PD ≤ 65 years.

Fig 1 represents the results of the follow-up study after training with dual-task activities in patients with Parkinson’s disease ≤ 65 years (N = 13), which was repeated a half year and one year later. Half year later the average reaction times on the first day were lower than that of the baseline, but only the reaction time of the task of ‘Simple’ was significant. The repetition of the dual-task activities after a half year led to a significant decrease in the average reaction time of ‘Bird’, ‘Divided’, and ‘Simple’. The columns represent the mean ± S.D: The black column indicates baseline values, the obliquely striped column shows the half year later, and the dotted column shows data one year later.

In our follow up study, improvement was maintained in the PD patients > 65 years in ‘Bird’ and ‘Simple’ as in the younger group of patients, although with higher significance (p < 0.001) (Fig 2). A similar change was observed in Hits (B: 60.3 ±21.6, H: 83.9 ± 26.1 p < 0.001 O: 89.4 ± 16.2 p < 0.001). The dual-task Habitat was not altered in PD patients ≤ 65 years, but results were significantly attenuated in PD patients > 65 years compared to the baseline (half year: p < 0.05, 12 months p < 0001) (Fig. 2). PD patients > 65 years was the only group where the Misses decreased after dual-task training for five days and this favorable tendency was observed 6 and 12 months later (B: 23.3 ±13.1 H: 16.8 ± 5.5 p < 0.05, O: 11.8 ± 6.0, p < 0.001).

**Fig 2.**
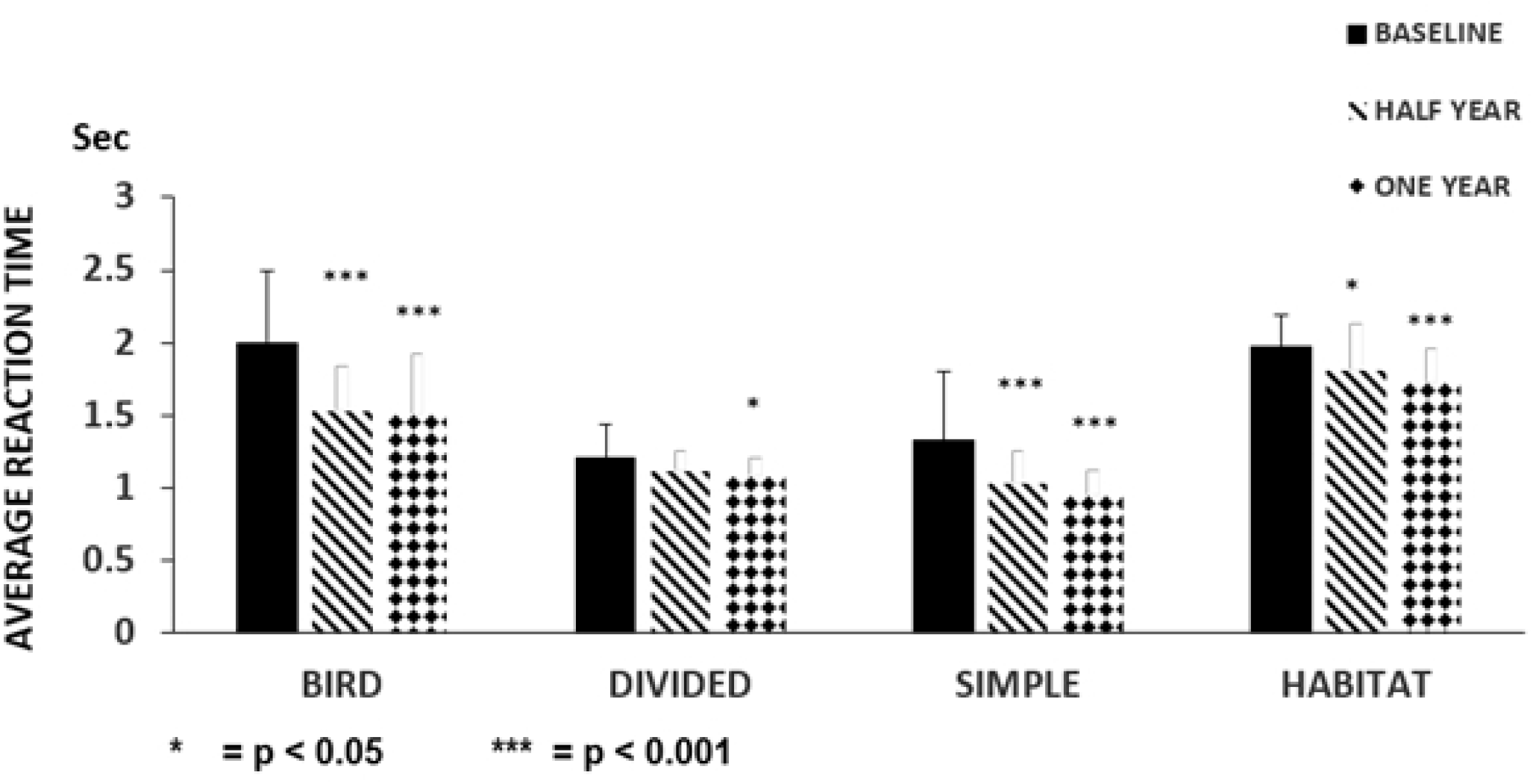
Follow-up of the effect of training with dual-task performances in PD > 65 years.

**Fig 2** depicts the results of the follow-up study after training with dual-task activities in the patients with Parkinson’s disease > 65 years (N = 13). Training for five days was repeated after one half and one year later. The average reaction times on the first days were compared with baseline results. A highly significant decrease was detected in dual-task activity ‘Bird’, ‘Simple’ and ‘Habitat’ after a half year, but not for ‘Divided’. The columns represent the mean ± S.D. The black column is for the baseline, the striped column is for half year later, and the dotted column is for one year later.

### Waking tests detecting lower limbs’ bradykinesia

During the dual task tests the participants used their lower limbs, so we tested their limb movements when walking as an objective assessment suitable for future comparisons. PD patients were compared to age-matched controls in different walking tests to detect their degree of bradykinesia. Patients under 65 years were significantly different from healthy controls during a walking test for 6 min (Control (C): 614 ± 125 m PD: 452 ± 143 m (p < 0.01) or along a 10 m distance (C: 5.8 ± 0.54 sec, PD: 8.3 ± 3.8 sec p < 0.01). Patients > 65 years tended to be slower, although not significantly, to the 6 min criterion: (C: 482 ± 97 m, PD: 392 ± 147 m), or walking 10 m: (C: 6.6 ± 1.6 sec, PD: 8.8 ± 2.9 sec p < 0.01). There were no differences between the first and fifth day of testing in the 6 min test, or between the 6 min and 10 m tests at 6 and 12 months after training.

### Cognitive tests

There were no significant differences in the scores of patients and control subjects in either age group (over or under 65 years) on the Mini Mental Rating Scale, the Ziehen-Ranschburg Word Pair Test, the Clock Drawing Test, the Hamilton Depression Scale, and the Trail Making Test.

## Discussion

This study has demonstrated the efficacy of dual-task tests in the assessment of cognitive decline in PD patients with H-Y I and II in an age-dependent way. The deterioration of dual-task performances was higher in the PD groups than in the age matched healthy controls, with the greatest decline in PD patients > 65 years. Training with dual-task performance tests for a short period had a rapidly beneficial effect on the delayed reaction times and the number of Hits. However, the values in patients never reached the level of the control participants. Dual-task training was most effective in patients > 65 years and cognitive improvements were retained for at least 6 months. The increased number of Misses may be the most sensitive parameter with which to assess cognitive decline. The simultaneous performance of two tasks induces a delay in the reaction time compared to single task activities, suggesting interference with attention and executive function in dual-task testing which are diminished in patients with PD [10, 11, 16, 25, 26]. A recent report noted a significant decline in the performance of treadmill walking with visuomotor and cognitive game dual-tasks [17], a conclusion well illustrated by Johansson et al. (2021).

Only a few previous studies have considered the influence of dual-task performance on cognitive function. Those studies mainly included walking tests, with a strong focus on the spatiotemporal parameters of walking [5, 6]. It has been reported that practice had an influence on the walking time component of a cognitive dual-task performance, while it had no effect on a motor dual-task activity [15]. In addition, the cognitive ability of PD patients was improved by cycling with cognitive tasks [27, 28]. Not only did training for several weeks have an impact on the motor outcome, but training with dual-tasks for one session also influenced the spatiotemporal walking parameters of PD patients [29]. Although attention has been drawn to the cognitive difference between the H-Y stage II and III [17], a later publication stressed the differences between PD patients with and without minimal cognitive impairment (MCI). These and other studies confirm that dual-task activities were strongly influenced by cognitive ability. In the light of these observations we have examined the number of Hits and Misses for the assessment of patients’ global cognitive ability, clearly showing differences between patients and age-matched healthy controls. Previous publications indicated the increased reaction time delays in PD patients, although at a later disease stage than the present study [13, 14, 16]. We have also highlighted the alterations in cognition assessed by dual-task tests between PD groups at different ages. The results are consistent with earlier work which studied an older population [30 - 32]. Our study indicates that cognitive deficits are detectable using dual tasks in patients younger than previously thought. It has been stated that cognitive impairment was apparent in 20 % of PD patients at the time of diagnosis [33], even when cognitive tests appeared to be within normal limits. The data were interpreted as indicating a subjective cognitive decline with no objective reasons [34]. This raises a critical issue in the rehabilitation of PD patients: there is an urgent need to detect cognitive impairment as early as possible, since it may predict future decline of PD patients [35]. At present detailed cognitive testing is not applied routinely to PD patients, as psychological tests of memory, executive function and attention are time consuming and need special requirements and dedicated staff. Dual-task performances can be performed relatively quickly and easily and can reveal much about the global cognition of patients. Here, a motor activity was employed as a part of the dual-task test, no difference was demonstrable between the motor and cognitive measurements.

Nevertheless, the parameters of every dual –task test slightly or significantly improved after training, mainly in PD patients over 65. We conclude that bradykinesia did not play a role in the improved results after dual task training, consistent with the work of Johansson et al. ⌠18⌡.

### Duration of training

Previous studies of dual task training have used extended periods of time, such as once or twice per week for several weeks to achieve improvement [4 - 6, 15, 36 -41⌡. One of our objectives was to determine whether shorter periods would be sufficient. One of our most striking observations was the rapidity of improvement after dual-task training. We hypothesize that rapid neural re-organisation led to decreased interaction between the dual tasks, a view which would underline the importance of cognition in dual-task tests, consistent with the poorer performances seen even in the early stages of PD. On the other hand, not every test in the dual-task paradigm was improved by training. This might suggest that neural network organization is related to the nature of the dual-task performances, with some tasks requiring a higher level of cognitive input. The idea would be consistent with the greater improvement after training in PD patients > 65 years. The greater the cognitive deficiency present, the more defective is the dual task performance (>65 years) and the greater the potential for improvement on even the minimal training used here. The present study confirmed that the delay observed in dual-task activity depends on the modality of the dual-task tests [42, 43]. Some dual task tests are better than others for generating cognitive improvements. One question to be addressed is whether a positive outcome in dual-tasks training transfers to other cognitive tasks. A few earlier studies of dual-task training focused on walking parameters, showing a slight, non-significant improvement in executive function assessed by the Trail Making Test in the DUALGAIT trial [5]. Evidence suggests that not only the trained dual tasks are improved, but also non-trained dual-tasks such as the auditory Stroop test (the DUALITY trial) [4]. Highly challenging tests led to greater improvement in cognitive performance, potentially transferrable to daily living activity [6]. The rapidity of cognitive improvement after training might indicate the general increase demanded in the exercise of attention, concentration and executive function necessary for the successful completion of a dual task session [7]. However, our observations suggested a diversion of attention onto irregularly changing objects. It is possible that, since our trial was conducted over several consecutive days, the novelty of those extraneous factors declined, allowing greater focus on the dual tasks.

### Repeat testing at 6 and 12 months

In the second part of this study, patients with PD were followed for one year to determine the sustainability of the response improvement. This represents the longest follow–up study using dual-task performances, since former studies generally followed the patients for only a few weeks after training [4 - 7, 40, 41]. The transferred effectiveness of dual-task training has been detected in in a few dual-task studies [6, 29]. Our extended successful testing at 12 months provides a strong argument that relatively transient training can give patients a prolonged period of enhanced cognitive function.

### Mechanisms

In spite of its effectiveness, the underlying central mechanisms of training by dual-task tests remain unclear. The complexity of motor and cognitive networks during dual-task activities were confirmed by the comparison of mental parameters with the interaction of dual-task tests, although there is little association between them [16]. An examination of functional networks relevant to dual-task activities were assessed by functional MRI, showing positive activity in the precuneus nuclei of the parietal lobes during dual-task activities compared to that of single tasks [44]. Furthermore, there was a shortage of activation in the right vermis of the cerebellum in PD patients, which may be responsible for the integration of motor and cognitive networks [45]. Some of these observations may be relevant to the present results.

Overall, this study indicates the usefulness of dual-task tests in the detection of global cognitive decay in the early stage of PD, where other assays are normal. The cognitive deficit apparent in dual-task testing was shortly overcome by dual-task training, but not to the level of controls. The cognitive improvement generated remained evidence for at least 6 months.

### Limitations

The main limitation of this study was that the separation of the primary task and the secondary task responses in these complex dual-task activities was not possible. Nevertheless our results may help understanding of the mechanisms by which training influences the overall interaction between the two components.

## Conclusion

Simultaneously performed dual task paradigms were used to monitor cognitive function in PD patients. Considering the response delay and the increased number of Misses in patients compared to controls, a decline in global cognition was demonstrated in Hoehn-Yahr stages I and II, mainly in patients above 65 years of age. Cognitive performance was improved by training with dual-task activities for only a few days, with greatest improvement in patients > 65 years. These favorable results were maintained for at least 6 months after the initial training. It is proposed that the use of dual-task testing for a few days should be considered as a highly recommended technique in the rehabilitation of patients with PD.

## Data Availability

All relevant data are within the manuscript and its Supporting Information files.

## Authors’ Contributions

Dalma Szögedi: execution of research project

Trevor W. Stone: Review and critique of the manuscript

Elek Dinya: Execution of statistical analysis

Judit Málly: conception of research project, writing of the first draft

## Authors’ declaration

We confirm that we have read the Journal’s position on issues involved in ethical publication and affirm that this work is consistent with those guidelines. All authors have substantially taken part in the study and the preparation of the manuscript. No undisclosed groups or persons have had a primary role in the study and/or in the manuscript preparation. All co-authors have read and approved the submission of the manuscript to the PLoS One. There is no ghost author among us. The work has not been published earlier nor is being considered for publication in another journal. The study was conducted in accordance with the Helsinki Declaration of 1975. This research received no specific grant from any funding agency in the public, commercial, or not-for-profit sectors.

## Supporting Information

**S1 Table 1 The demographic data of the comparative study with dual-task tests**

**S2 Table 2 Demographic data of patients with Parkinson’s Disease included in this follow-up study**.

**S3 Table 3 Comparing the patients’ and controls’ results in the dual-task activity during training with dual-task tests ≤ 65 years**.

Table 3 shows the results of the dual-task performances of Parkinsonian patients ≤ 65 years compared with that of the controls. Data of Bird, Simple, Divided, Habitat are given in seconds of their mean ± S.D. Hits and Misses are given in the mean ± S.D.

**S4 Table 4 Comparing the patients’ and controls’ results in the dual-task activity during training with dual-task tests > 65 years**.

**S1 Fig 1 Follow-up of the effect of training with dual-task performances in the PD**

**S2 Fig 2. Follow-up of the effect of training with dual-task performances for five days in patients with PD > 65 years**

Fig 2 depicts the results of the follow-up study after training with dual-task activities in the patients with Parkinson’s disease > 65 years (N = 13). Training for five days was repeated after one half and one year later. The average reaction times on the first days were compared with baseline results. A highly significant decrease was detected in dual-task activity ‘Bird’, ‘Simple’ and ‘Habitat’ after a half year, but not for ‘Divided’. The columns represent the mean ± S.D. The black column is for the baseline, the striped column is for half year later, and the dotted column is for one year later.

## References

1. Leung IH, Walron CC, Hallock H, Lewis SJ, Valenzuela M, Lampit A. Cognitive training in Parkinson disease: A systematic review and meta-analysis. Neurology. 2015; 85(21):1843–1851. https://doi.org/10.1212/WNL.0000000000002145 PMID: 26519540

2. Hindle JV, Petrelli A, Clare L, Kalbe E. Nonpharmacoological enhancement of cognitive function in Parkinson’s disease: A systematic review. Mov Disord. 2013; 28:1034–1049. https://doi.org/10.1002/mds.25377 PMID: 23426759

3. David FJ, Robichaud JA, Leurgans SE, Poon C, Kohrt WM, Goldman JG, et al. Exercise improves cognition in Parkinson’s disease: the PRET-PD randomized clinical trial. Mov Disord. 2015; 30(12):1657–1663. https://doi.org/10.1002/mds.26291 PMID: 26148003

4. Strouwen C, Molenaar EALM, Münks L, Keus SHJ, Zijlmans JCM, Vandenberghe W, et al. Training dual tasks together or apart in Parkinson’s disease: Results from the DUALITY trial. Mov Disord. 2017; 32(8):1201–1210. https://doi.org/10.1002/mds.27014 PMID: 28440888

5. San Martín Valenzuela C, Moscardó LD, López-Pascual J, Serra-Añó P, Tomás JM. Effects of dual-task group training on gait, cognitive executive function, and quality of life in people with Parkinson’s disease: Results of randomized controlled DUALGAIT trial. Arch Phys Med Rehabil. 2020; 101(11):1849–1856. https://doi.org/10.1016/j.apmr.2020.07.008 PMID: 32795562

6. Conradsson D, Löfgren N, Nero H, Hangströmer M, Stähle A, Lökk J. et al. The effects of highly challenging balance training in elderly with Parkinson’s disease: A randomized controlled trial. Neurorehabil Neural Repair. 2015; 29(9):827–836. https://doi.org/10.1177/1545968314567150 PMID: 25608520

7. Beck EN, Intzandt BN, Almeida QJ. Can dual-task walking improve in Parkinson’s disease after external focus of attention exercise? A single blind randomized controlled trial. Neurorehab Neurol Repair. 2018; 32(1):18–33. https://doi.org/10.1177/1545968317746782 Epub 2017 Dec 20. PMID: 29262749

8. Schwab RS, Chafetz ME, Walker S. Control of two simultaneous voluntary motor acts in normals and in Parkinsonism. AMA Arch Neurol Psychiatry. 1954; 72(5):591–598. doi: https://doi.org/10.1001/archneurpsyc.1954.02330050061010 PMID: 13206480

9. Benecke R, Rothwell JC, Dick JPR, Day BL, Marsden CD. Performance of simultaneous movements in patients with Parkinson’s disease. Brain. 1986; 109(Pt 4):739–757. https://doi.org/10.1093/brain/109.4.739 PMID: 3730813

10. Benecke R, Rothwell JC, Dick JPR, Day BL, Marsden CD. Simple and complex movements off and on treatment in patients with Parkinson’s disease. J. Neurol. Neurosurg Psychiatry. 1987; 50(3):296–303. https://doi.org/10.1136/jnnp.50.3.296 PMID: 3559611

11. Brown RG, Marsden CD. Dual task performance and processing resources in normal subjects and patients with Parkinson’s disease. Brain. 1991; 114(Pt 1A):215–231. https://doi.org/10.1093/oxfordjournals.brain.a101858 PMID: 1998883

12. Plotnik M, Dagan Y, Gurevich T, Giladi N, Hausdorff JM. Effects of cognitive function on gait and dual tasking abilities in patients with Parkinson’s disease suffering from motor response fluctuations. Exp Brain Res. 2011; 208(2):169–179. https://doi.org/10.1007/s00221-010-2469-y Epub 2010 Nov 10. PMID: 21063692

13. Salazar RD, Ren X, Ellis TD, Toraif N, Barthelemy OJ, Neargarder S, et al. Dual tasking in Parkinson’s disease: Cognitive consequences while walking. Neuropsychology. 2017; 31(6):613–623. https://doi.org/10.1037/neu0000331 PMID: 28414497

14. Raffegeau TE, Krehbiel LM, Kang N, Thijs FJ, Altmann LJP, Cauraugh JH, et al. A meta-analysis: Parkinson’s disease and dual-task walking. Parkinsonism Relat Disord. 2019; 62:28–35. https://doi.org/10.1016/j.parkreldis.2018.12.012 Epub 2018 Dec 12. PMID: 30594454

15. Yang YR, Cheng SJ, Lee YJ, Liu YC, Wang RY. Cognitive and motor dual task gait training exerted specific training effects on dual task gait performance in individuals with Parkinson’s disease: A randomized controlled pilot study. PLoS One. 2019; 14(6):1–7. https://doi.org/10.1371/journal.pone.0218180 PMID:31220121

16. Dalrymple-Alford JC, Kalders AS, Jones RD, Watson RW. A central executive deficit in patients with Parkinson’s disease. J Neurol Neurosurg Psychiatry. 1994; 57(3):360–7. https://doi.org/10.1136/jnnp.57.3.360 PMID: 8158188;

17. Bhatt M, Mahana B, Ko JH, Kolesar TA, Kanitkar A, Szturm T. Computerized dual-task testing of gait visuomotor and cognitive functions in Parkinson’s disease: Test-retest reliability and validity. Front Hum Neurosci. 2021; 15:706230. https://doi.org/10.3389/fnhum.2021.706230 PMID: 34335213

18. Johansson H, Ekman U, Rennie L, Peterson DS, Leavy B, Franzén E. Dual-task effects during a motor-cognitive task in Parkinson’s disease: Patterns of Prioritization and the influence of cognitive status. Neurorehab Neural Repair. 2021; 35:356–366. https://doi.org/10.1177/1545968321999053 PMID: 33719728

19. Folstein MF, Folstein SE, McHugh PR. “Mini-mental state”. "Mini-Mental State” A practical method for grading the cognitive state of patients for the clinician. J Psychiatr Res. 1975; 12(3):189–98. https://doi.org/10.1016/0022-3956(75)90026-6 PMID: 1202204.

20. Reitan RM. Trail Making Test: Manual for administration and scoring. Reitan Neuropsychology Laboratory: Tempe, AZ:USA 1992; Corpus ID:141448957

21. Sunderland T, Hill JL, Mellow AM, Lawlor BA, Gundersheimer J, Newhouse PA, et al. Clock drawing in Alzheimer’s disease: a novel measure of dementia severity. J Am Geriatr Soc. 1989; 37(8):725–729. https://doi.org/10.1111/j.1532-5415.1989.tb02233.x PMID: 2754157

22. Hamilton M. A rating scale for depression. J. Neurol. Neurosurg Psychiatry. 1960; 23(1):56–62. http://dx.doi.org/10.1136/jnnp.23.1.56 PMID: 14399272

23. Hoehn MM, Yahr MD. Parkinsonism: onset, progression, and mortality. Neurology. 1967; 17(5):427–442. https://doi.org/10.1212/WNL.17.5.427 PMID: 6067254

24. Fahn S, Elton R. Members of the UPDRS Development Committee. 1987. in: Fahn S, Marsden CD, Calne DR, Goldstein M. eds. Recent Developments in Parkinson’s Disease, Florham Park, NJ: Macmillan Health Care Information. 1987; 2:153–163; 293–304.

25. Woodward TS, Bub DN, Hunter MA. Task switching deficits associated with Parkinson’s disease reflect depleted attentional resources. Neurospsychologia. 2002; 40(12):1948–1955. https://doi.org/10.1016/S0028-3932(02)00068-4 PMID: 12207992

26. Li Z, Wang T, Liu H, Jiang Y, Wang Z, Zhuang J. Dual-task training on gait, motor symptoms, and balance in patients with Parkinson’s disease: a systematic review and meta-analysis. Clin Rehabil. 2020; 34(11):1355–1367. https://doi.org/10.1177/0269215520941142 PMID: 32660265

27. Altmann LJP, Stegemöller E, Hazamy AA, Wilson JP, Okun MS, McFarland NR, et al. Unexpected dual task benefits on cycling in Parkinson disease and healthy adults: A neuro-behavioral model. PLoS One. 2015; 10(5):e0125470. https://doi.org/10.1371/journal.pone.0125470 PMID: 25970607

28. Hsiu-Chen C, Chiung-Chu C, Jiunn-Woei L, Wei-Da C, Yi-Hsin W, Ya-Ju C, et al. The effects of dual-task in patients with Parkinson’s disease performing cognitive-motor paradigms. J Clin Neurosci. 2020; 72:72–78. https://doi.org/10.1016/j.jocn.2020.01.024 PMID: 31952973

29. Brauer SG, Morris ME. Can people with Parkinson’s disease improve dual tasking when walking? Gait Posture. 2010; 31(2):229–233. https://doi.org/10.1016/j.gaitpost.2009.10.011 Epub 2009 Dec 6. PMID: 19969461

30. Malcolm BR, Foxe JJ, Butler JS, De Sanctis P. The aging brain shows less flexible reallocation of cognitive resources during dual-task walking: a mobile brain/body imaging (MoBI) study. Neuroimage. 2015; 117:230–242. https://doi.org/10.1016/j.neuroimage.2015.05.028 PMID: 25988225

31. Brustio PR, Magistro D, Zecca M, Rabaglietti E, Liubicich ME. Age-related decrements in dual-task performance: Comparison of different mobility and cognitive tasks. A cross sectional study. A cross sectional study. PLoS One. 2017; 12(7):e0181698. https://doi.org/10.1371/journal.pone.0181698 PMID: 28732080

32. Ehsani H, Mohler MJ, O’Connor K, Zamrini E, Tirambulo C, Toosizadeh N. The association between cognition and dual-tasking among older adults: the effect of motor function type and cognition task difficulty. Clin Interv Aging. 2019; 14:659–669. https://doi.org/10.2147/CIA.S198697 PMID: 31040655

33. Zhang Q, Aldridge GM, Naravanan NS, Anderson SW, UC EY. Approach to cognitive impairment in Parkinson’s disease. Neurotherapeutics. 2020; 17(4):1495–1510. https://doi.org/10.1007/s13311-020-00963-x PMID: 33205381

34. Aarsland D, Creese B, Politis M, Chaudhuri KR, Ffytche DH, Weintraub D, et al. Cognitive decline in Parkinson disease. Nat Rev Neurol. 2017; 13(4):217–231. https://doi.org/10.1038/nrneurol.2017.27 PMID: 28257128

35. Burn D, Weintraub D, Robbins T. Introduction: The importance of cognition in movement disorders. Mov Disord. 2014; 29(5): 581–583. https://doi.org/10.1002/mds.25871 PMID: 24757107

36. Soliveri P, Brown RG, Jahanshahi M, Marsden CD. Effect of practice on performance of a skilled motor task in patients with Parkinson’s disease. J Neurol Neurosurg Psychiatry. 1992; 55(6):454–460. http://dx.doi.org/10.1136/jnnp.55.6.454 PMID: 1619411

37. Van Selst M, Ruthruff E, Johnston JC. Can practice eliminate the psychological refractory period effect? J Exp Psychol Hum Percept Perform. 1999; 25(5):1268–1283. https://doi.org/10.1037/0096-1523.25.5.1268 PMID: 10531663

38. Ruthruff E, Johnston JC, Van Selst M. Why practice reduces dual-task interference. J Exp Psychol Hum Percept Perform. 2001; 27(1):3–21. https://doi.org/10.1037/0096-1523.27.1.3 PMID: 11248938

39. Fritz NE, Cheek FM, Nichols-Larsen DS. Motor-cognitive dual-task training in persons with neurologic disorders: A systematic review. J. Neurol. Phys. Ther. 2015; 39(3):142–153. https://doi.org/10.1097/NPT.0000000000000090 PMID: 26079569

40. Geroin C, Nonnekes J, de Vries NM, Strouwen C, Smania N, Tinazzi M, et al. Does dual-task training improve spatiotemporal gait parameters in Parkinson’s disease? Parkinsonism Relat. Disord. 2018; 55:86–91. https://doi.org/10.1016/j.parkreldis.2018.05.018 PMID: 29802080

41. Wollesen B, Rudnik S, Gulberti A, Cordes T, Gerloff C, Poetter-Nerger M. A feasibility study of dual-task strategy training to improve gait performance in patients with Parkinson’s disease. Sci Rep. 2021; 11(1):12416. https://doi.org/10.1038/s41598-021-91858-0 PMID: 34127721

42. Hazeltine E, Ruthruff E, Remington RW. The role of input and output modality pairings in dual-task performance: Evidence for content-dependent central interference. Cogn Psychol. 2006; 52(4):291–345. https://doi.org/10.1016/j.cogpsych.2005.11.001 PMID: 16581054

43. Halvorson KM, Hazeltine E. Separation of tasks into distinct domains, not set-level compatibility, minimizes dual-task interference. Front Psychol. 2018; 10:711. https://doi.org/10.3389/fpsyg.2019.00711 PMID: 30984091

44. Wu T, Hallett M. Neural correlates of dual task performance in patients with Parkinson’s disease. J Neurol Neurosurg Psychiatry. 2008; 79(7):760–766. https://doi.org/10.1136/jnnp.2007.126599 Epub 2007 Nov 15. PMID: 18006652

45. Gao L, Zhang J, Hou Y, Hallett M, Chan P, Wu T. The cerebellum in dual-task performance in Parkinson’s disease. Sci. Rep. 2017; 7:45662 https://doi.org/10.1038/srep45662 PMID: 28358358

